# Prevalence, pattern and caregiver responses to adverse events following immunisation in selected secondary and tertiary hospitals in Osogbo Nigeria: A Cross-Sectional Study

**DOI:** 10.64898/2026.09.15.26363153

**Authors:** Funso Abidemi Olagunju, Sunday Charles Adeyemo, Abimbola Ololade Odeyemi, Efeturi Agelebe, Abisola Oluwatoyin Omoboyeje, Kehinde Awodele, Rukayat Oyelami-Adegbite, Samuel Olorunyomi Oninla, Eniola Dorcas Olabode

**Author notes:** Corresponding author: Sunday Charles Adeyemo Institut Superieur de Sante, Niamey, Niger Republic.

## Abstract

**Background:** Surveillance of adverse events following immunization (AEFI) and appropriate reporting are essential ingredients of an effective immunization programme. Despite increasing recognition by caregivers, the response to it remains a major problem, with the majority being inappropriate. This study is therefore intended to determine the prevalence, pattern, and determinants of caregivers’ response regarding AEFI in Nigerian hospitals.

**Methodology:** A descriptive cross-sectional study was conducted among 368 caregiver-infant pairs across 3 Nigerian hospitals, using semi-structured questionnaires to document sociodemographic characteristics and the prevalence, pattern, and determinants of caregivers’ response. Statistical analyses were performed using SPSS version 27. Predictors of AEFI and determinants of caregivers’ response were assessed using Pearson’s chi-square and logistic regression. A p-value < 0.05 was considered statistically significant.

**Results:** The prevalence of AEFI was 71.7%, with inconsolable crying/irritability, fever, and swelling at the vaccination site as the leading cases. Infant age, weight, and gestational status at birth were predictors of AEFI occurrence. Only 8 (3.0%) caregivers presented at a healthcare facility when AEFI was noticed (appropriate response), while 360 (97.0%) did not (inappropriate). The relationship of the caregiver to the vaccinated infant (mother) was found to be a determinant of caregivers’ response to the occurrence of AEFI.

**Conclusion:** An increasing prevalence of AEFI that was not followed by an optimally appropriate response from caregivers has been demonstrated. This calls for more public enlightenment focusing on the need to report AEFI at healthcare facilities, as this will help in ensuring vaccine safety and improvement in the immunization programme.

## Introduction

Adverse events following immunization (AEFI) are unfavourable medical conditions of concern that do not necessarily have a causal association with the vaccines.^1,2^ They are often experienced by infants and relayed by caregivers following the receipt of the vaccines. They impact the global vaccination programme through loss of trust in the programme, resulting in an increased tendency for vaccine rejection, increased dropout rates, and the overall consequence is an epidemic of vaccine-preventable diseases.^2,3^

World Health Organization (WHO) classifies AEFI based on cause-specificity into vaccine reactions (product-related and quality defect-related), immunization error-related reactions, immunization anxiety-related reactions, and coincidental events.^4,5^ Vaccine product–related reactions occur because of some intrinsic characteristics of the vaccine or its constituents, while vaccine quality defect–related reactions are due to challenges regarding the quality during production, storage, and how vaccines are conveyed to the final consumers (cold chain system) that affect the effectiveness of the vaccines. They include minor ones such as pain, swelling at the site of injection, fever, malaise, and severe ones such as seizures.^4,5^

Improper handling and dispensing techniques, which may result in possible contamination, inappropriate route of administration, and dosage, are hallmarks of immunization error–related reactions.^4,5^ They contributed the largest share to AEFI cases. Possible cases of AEFI due to immunization errors include injection site swelling, cellulitis and abscess arising from reuse of disposable syringes or needles, and sciatic nerve injury arising from wrong route of administration.^4,5^

Immunization anxiety–related reactions are psychosomatic manifestations that have no true link to the vaccine product, compromise with the quality of the vaccine, or a blunder of the immunization programme.^4,5^ They are associated with anxiety, and they include hyperventilation-mediated reactions, stress-related mental and behavioural events, and vasovagal-mediated disorders. Coincidental events are a series of medical complaints that occur after a particular vaccine has been received but whose causal relationship cannot be established to it or its administration. They are inevitable occurrences, and diligent understanding of the usual incidence of such complaints or conditions in relation to the timing and coverage of immunization helps to have a grasp of the number of coincidental events that follow the immunization.^4,5^

The prevalence of AEFI varies across regions. In 2015, 55% of countries in Europe reported at least 10 AEFI per 100,000 surviving infants, 43% in the Eastern Mediterranean Region, 33% in the Western Pacific Region, 60% in the Americas, 21% in the African Region, and 27% in the South-East Asia Region.^6,7^ In Nigeria, the prevalence of AEFI has ranged from 19.3% to 57%.^2^ Regarding patterns, Mathew et al.^8^ in India reported fever as the most common manifestation. Ogundele et al.^2^ in Ile-Ife, Southwest Nigeria, also reported fever (88%) and swelling at the injection site (78%) as the leading AEFI in their study. Similarly, Maizare et al.^9^ in Kano, Northern Nigeria, reported fever (66.4%) and persistent inconsolable cry (38.4%) as the most common AEFI. In another facility-based study from Rivers State in South-South Nigeria,^10^ the most common AEFI were fever (84.1%) and swelling at the injection site (45.8%).

Upon observing an AEFI, caregivers must approach a healthcare facility and report it. Appropriate reporting of AEFI remains an essential part of vaccine safety surveillance, which fosters prompt identification of potential risks, informs public health decisions, improves immunization programs, and ensures public confidence.^11,12^ Varying reports of caregivers’ appropriate responses (14%; Jos, 42.3%; Ile-Ife, 99%; Benin City) have been documented across Nigeria, with the majority being suboptimal.^2,13^

It is clear from the foregoing that there are gaps between documenting the prevalence of AEFI and instituting an appropriate response (presenting to a healthcare facility) by caregivers when AEFI are noticed. These gaps need to be addressed with a view to proffering strategic interventions that will translate into appropriate actions whenever the occurrence of AEFI is established. This multi-Center study, therefore, is intended to determine the prevalence, pattern, and determinants of caregivers’ response regarding AEFI in Nigerian hospitals.

## METHODOLOGY

### Study setting

The study was conducted at Osun State University (UNIOSUN) Teaching Hospital, State Specialists’ Hospital Asubiaro (SSHA), and Our Lady of Fatima Catholic Hospital (OLFCH), all in Osogbo, the State Capital of Osun State. The study was carried out at the immunization centres of the hospitals. On average, about 150 to 200 infants are vaccinated in a week at UNIOSUN Teaching Hospital, 250 to 300 infants are vaccinated at the State Specialists’ Hospital, while at Our Lady of Fatima Catholic Hospital, 50 to 80 infants are vaccinated weekly. Nurses and community health extension workers (CHEWS) are usually the healthcare workers at the centres. The services rendered at the centre include vaccination of newborns, infants, older children, and adults. Others include health education and promotion on topics such as immunization, breastfeeding and infant nutrition, and family planning for mothers.

### Study population

The study population was caregiver-infant pairs who presented for vaccination at the immunization centres of the hospitals. The target population was caregivers and infants who had had at least the first routine vaccination schedule, which is Bacillus Calmette-Guérin (BCG), Oral polio vaccine (OPV), and Hepatitis B vaccine (HBV).

### Inclusion and exclusion criteria

#### Inclusion criteria

Caregivers whose infants/children have had at least the first routine vaccination schedule (BCG, OPV, and HBV) and gave informed consent to participate in the study.

#### Exclusion criterion

Caregivers who were mentally indisposed to participate in the study and infants who were ill were excluded from the study

### Study duration

The study was conducted over 4 months, from October 2025 to February 2026.

### Study design

The study deployed a cross-sectional design.

### Sample size determination

The sample was determined using Leslie-Fisher’s formula.

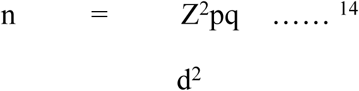

where n = sample size

z = Standard normal deviation at the required confidence level of 1.96

p = Proportion in the characteristics being measured (p = 33.1%; prevalence from a previous study) ^15^

q = 1 – p

d = Level of statistical significance set (i.e., 0.05)

n = 1.96^2^ x 0.331 (1-0.331) 0.0025 = 340

The minimum sample size was estimated to be 340. 10% of the calculated 340 was added to make up for non-response. Hence, the total number of participants was 374.

### Sampling techniques

The study utilized a multistage sampling technique as follows:

Stage One: Proportionate sampling, which was deployed to ensure each facility had a representative share of the total sample size based on the weekly vaccination uptake by caregiver-infant pairs. Using the ratio 1:2:3 (80, 150, 300 vaccination uptakes from OLFCH, UTH, AND SSHA, respectively), the total number of participants from each facility amounted to 62, 125, and 187 for OLFCH, UTH, and SSHA, respectively.

Stage Two: Systematic random sampling was used to select the participants in each of the facilities. A list of the caregiver-infant pairs who presented for vaccination was collated to determine the total number for the day (sampling frame). Thereafter, the sampling intervals were determined by dividing the total/overall sample size by the sample population size from each facility (OLFCH =374/62 = 6, UTH = 374/125 = 3, and SSHA = 374/187 = 2).

Stage Three: Simple random sampling was used to select the first caregiver-infant pairs, and the others were selected using the sampling interval calculated for each facility from the sampling frame till the total sample for each facility was reached.

### Study instrument

The study instrument was a self-administered, semi-structured questionnaire. The questionnaire consisted of 3 sections. The first section contained socio-demographic information of the caregiver-infant pair. Their socioeconomic class was determined using Ibadin et al,’^16^ socioeconomic class. The second section contained a list of AEFI medical conditions, which the caregiver-infant pairs responded to by ticking the appropriate one as experienced by them. The third section comprised a question which was asked to determine the response of the caregiver-infant pairs to the experience they had with AEFI. The response was judged as appropriate when caregiver-infant pairs approached healthcare facilities such as hospitals and PHCs for management of the AEFI, and it was inappropriate if they did not.

The questionnaire was pretested at the Immunization Centre of Cottage Hospital Ede, Osun State, Nigeria using 10% of the calculated sample size. Clarity and validity of the content of the questionnaire were ensured by expert review by Consultants in Public Health and Paediatric and Child Health. Internal consistency was also ensured by obtaining Cronbach’s alpha above 0.7.

### Data collection

An informed consent form, which explained the purpose of the study, was given to all the caregiver-infant pairs to go through. The purpose of the study was emphasized; they were made to understand that their participation was voluntary and that they could withdraw without any consequence, and the information they provided was treated with utmost confidentiality. Thereafter, they were offered the form, and written consent was obtained. Two research assistants (per facility) who were doctors were trained, and they assisted with the collection of the information needed as reflected in the questionnaire. All information obtained from each participant was kept on a computer with a password known to the researchers only.

### Data management and analysis

Questionnaires were manually sorted out for errors and omissions at the end of the data collection. The data were analyzed using Statistical Package for Social Sciences (SPSS) version 27.0 (SPSS, Chicago Inc, IL, USA). Categorical variables such as socioeconomic class of the caregivers-infants’ pairs, sex, age categories, and patterns of AEFI were summarized using proportions and percentages. Continuous variables such as age and weight of the participants were summarized using the mean ± standard deviation (SD) for normally distributed variables. The determinants of caregivers-infants’ pairs’ response to AEFI were determined using bivariate analysis (Chi-square test) and multivariate analysis (binomial regression). Statistical significance was set at a p-value less than 0.05 for all values of test statistics.

### Ethical approval

All ethical principles guiding the conduct of research, such as informed consent, beneficence, non-maleficence, confidentiality, justice, autonomy, etc., were strictly adhered to. Ethical approval was obtained from the Osun State Health Research Ethical Committee with protocol number OSHREC/PRS/569T/1279.

## Results

### Socio-demographic characteristics of the caregiver-infant pairs

Three hundred and seventy-four (374) caregiver-infant pairs participated in the study. Out of these, 368 (98.4%) questionnaires were properly filled, while 6 (1.6%) were not filled properly. Hence, data analysis was carried out on the 368 properly filled questionnaires. Mothers constituted the largest percentage (358; 97.3%) of the caregivers, while among the infants, sex distribution was almost equal. More than three-quarters (313; 85%) resided within Osogbo, while 55 (14.9%) lived outside Osogbo. The mean age and standard deviation of the caregivers were 30.0 ± 4.8 years; those of the infants were 6.4 ± 4.7 months. The mean weight and standard deviation of the infants were 6.5 ± 2.1Kg. Other details are as shown in Tables 1 and 2.

**Table 1:** Socio-demographic characteristics of the caregivers (N = 368)

| **Variables** | **Number (N=368)** | **Percentage (%)** |
| --- | --- | --- |
| **Age groups (Years)** |  |  |
| 11-20 | 4 | 1.1 |
| 21-30 | 214 | 58.2 |
| 31-40 | 143 | 38.9 |
| 41-50 | 7 | 1.9 |
| *Total* | *368* | *100.0* |
| **Caregivers' relationship to infants** |  |  |
| Mother | 358 | 97.3 |
| Aunt | 7 | 1.9 |
| Father | 3 | 0.9 |
| *Total* | *368* | *100.0* |
| **Educational status** |  |  |
| Primary | 38 | 10.3 |
| Secondary | 176 | 47.8 |
| Tertiary | 149 | 40.5 |
| Informal | 5 | 1.4 |
| *Total* | *368* | *100.0* |
| **Occupational status** |  |  |
| Civil servants | 88 | 23.9 |
| Artisan | 73 | 19.8 |
| Trading | 192 | 52.2 |
| Farming | 1 | 0.3 |
| Unemployed | 14 | 3.8 |
| *Total* | *368* | *100.0* |
| **Socio-economic class** |  |  |
| Low | 148 | 40.2 |
| Middle | 158 | 42.9 |
| High/Upper | 62 | 16.9 |
| *Total* | *368* | *100.0* |

**Table 2:** Infants’ demographic and health-related characteristics (N = 368)

| **Variables** | **Number (N=368)** | **Percentage (%)** |
| --- | --- | --- |
| **Age groups (Months)** |  |  |
| 1 – 5 | 174 | 47.3 |
| 6 – 10 | 124 | 33.7 |
| > 10 | 70 | 19.0 |
| *Total* | *368* | *100.0* |
| **Weight groups (Kg)** |  |  |
| 2 – 3.9 | 29 | 7.9 |
| 4 – 5.9 | 132 | 35.9 |
| 6 – 7.9 | 112 | 30.4 |
| ≥ 8 | 95 | 25.8 |
| *Total* | *368* | *100.0* |
| **Sex** |  |  |
| Male | 183 | 49.7 |
| Female | 185 | 50.3 |
| *Total* | *368* | *100.0* |
| **Mode of delivery** |  |  |
| Caesarean section (CS) | 102 | 27.7 |
| Spontaneous vertex delivery (SVD) | 266 | 72.3 |
| *Total* | *368* | *100.0* |
| **Gestation status at delivery** |  |  |
| Term | 348 | 94.6 |
| Preterm | 20 | 5.4 |
| *Total* | *368* | *100.0* |

### Prevalence, patterns of AEFI cases, implicated vaccines and schedules, and predictors of occurrence among the infants

Out of the 368 infants that participated in the study, 264 (71.7%) experienced AEFI while 104 (28.3%) did not. Regarding the patterns of the AEFI cases, irritability / excessive crying, fever, and painful swelling at the site of vaccine administration were the leading cases (Table 3). The vaccine schedules found to be associated with many occurrences of AEFI were those that the infants had at birth, 6^th^, 10^th^, and 14^th^ week (Table 4). Bivariate analysis revealed age of the infant (p = 0.024), weight (p = 0.024), and gestational status at birth (p = 0.006) as predictors of AEFI occurrence (Table 5).

**Table 3:** Patterns of AEFI cases.

| **Patterns of AEFI cases** | **Number and percentages of AEFI cases**<br>**n (%) #** |
| --- | --- |
| Irritability / Excessive crying | 169 (39.9) |
| Fever | 134 (31.7) |
| Painful swelling at the site of vaccine administration | 109 (25.8) |
| Body itching (Anaphylaxis) | 7 (1.7) |
| Convulsion | 3 (0.7) |
| Bleeding | 1 (0.2) |
| *Total* | *423 (100.0)* |

**Table 4:** AEFI occurrences and implicated vaccine schedules.

| **Vaccine schedules** | **Vaccines** | **Number of AEFI occurrences**<br>**n / %** |
| --- | --- | --- |
| At birth | BCG, OPV <sup>0</sup>, HBV | 49 (18.6) |
| 6 weeks | Pentavalent <sup>1</sup>, PCV <sup>1</sup>, Rotavirus <sup>1</sup>, OPV <sup>1</sup> | 104 (39.4) |
| 10 weeks | Pentavalent <sup>2</sup>, PCV <sup>2</sup>, Rotavirus <sup>2</sup>, OPV <sup>2</sup> | 56 (21.2) |
| 14 weeks | Pentavalent <sup>3</sup>, PCV <sup>3</sup>, OPV <sup>3</sup>, IPV | 42 (15.9) |
| 6 months | Vitamin A (First dose) | 11 (4.2) |
| 9 months | Measles, Yellow fever | 2 (0.7) |
| 12 months | Vitamin A (Second dose) | 0 (0.0) |
| *Total* |  | *264 (100.0)* |

**Table 5:** Predictors of occurrence of AEFI among the infants (Bivariate analysis)

| **Variables** | **AEFI: Yes**<br>**n (%)** | **AEFI: No**<br>**n (%)** | **Test statistics**<br>**$\chi^2$** | **p-value** |
| --- | --- | --- | --- | --- |
| **Age groups**<br>**(months)** |  |  |  |  |
| 1 – 5 | 129 (48.9) | 45 (43.3) | 7.492 | **0.024** |
| 6 – 10 | 94 (35.6) | 30 (28.8) |  |  |
| ≥ 10 | 41 (15.5) | 29 (27.9) |  |  |
| Total | 264 (100.0) | 104 (100.0) |  |  |
| **Sex** |  |  |  |  |
| Male | 129 (48.9) | 54 (51.9) | 0.279 | 0.597 |
| Female | 135 (51.1) | 50 (48.1) |  |  |
| Total | 264 (100.0) | 104 (100.0) |  |  |
| **Weight groups**<br>**(Kg)** |  |  |  |  |
| 2 – 3.9 | 21 (8.0) | 8 (7.6) | 9.464 | **0.024** |
| 4 – 5.9 | 95 (36.0) | 37 (35.6) |  |  |
| 6 – 7.9 | 90 (34.1) | 22 (21.2) |  |  |
| ≥ 8 | 58 (21.9) | 37 (35.6) |  |  |
| Total | 264 (100.0) | 104 (100.0) |  |  |
| **Gestational**<br>**status at birth** |  |  |  |  |
| Term | 255 (96.6) | 93 (89.4) | 7.459 | **0.006** |
| Preterm | 9 (3.4) | 11 (10.6) |  |  |
| Total | 264 (100.0) | 104 (100.0) |  |  |
| **Mode of**<br>**delivery** |  |  |  |  |
| Caesarean section (CS) | 80 (30.3) | 22 (21.2) | 3.117 | 0.077 |
| Spontaneous vertex delivery (SVD) | 184 (69.7) | 82 (78.8) |  |  |
| Total | 264 (100.0) | 104 (100.0) |  |  |

$\chi^2$ = Pearson's Chi-square. \* = Significant

### Caregivers’ response to occurrence of AEFI and associated determinants

Out of the 264 caregivers who noticed AEFI in their infants, more than two-thirds [196; (74.2%)] resorted to self-medications ranging from administration of paracetamol drops, ibuprofen syrup, to hot and cold massaging of injection sites (inappropriate response). Sixty (22.8%) caregivers did not do anything (inappropriate response); only 8 (3.0%) did present their infants to the healthcare facility for review and proper management (appropriate response). The overall responses of the caregivers are as shown in Figure 1.

**Figure 1:**
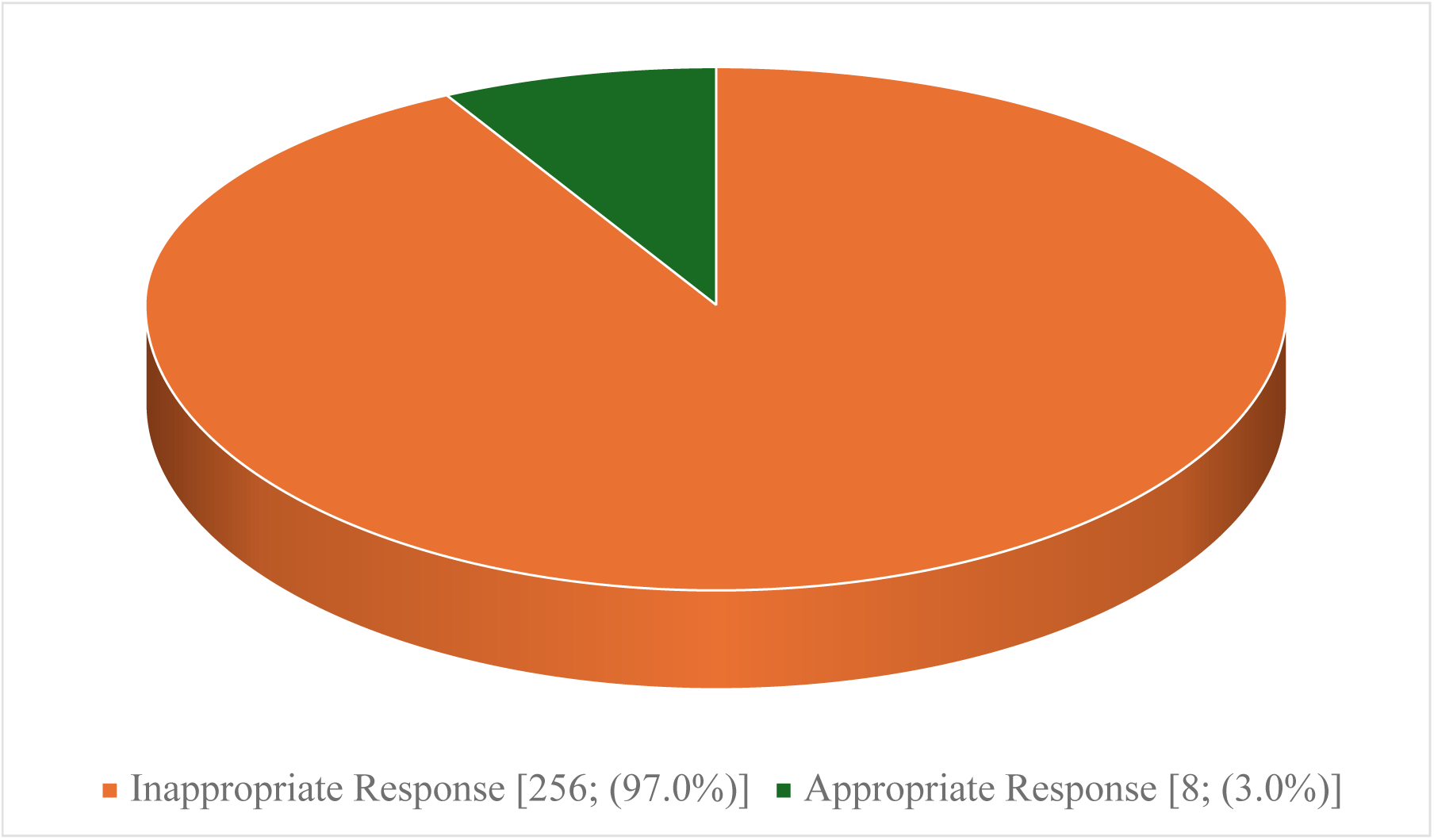
Caregivers’ response to occurrence of AEFI.

### Determinants of Caregivers’ Response to Occurrence of AEFI

Bivariate analysis using Pearson’s Chi-square revealed that sex of the caregivers (p = 0.002) and their relationship to the vaccinated infants (p = 0.001) were determinants of the caregivers’ response to the occurrence of AEFI (Table 6).

**Table 6:**
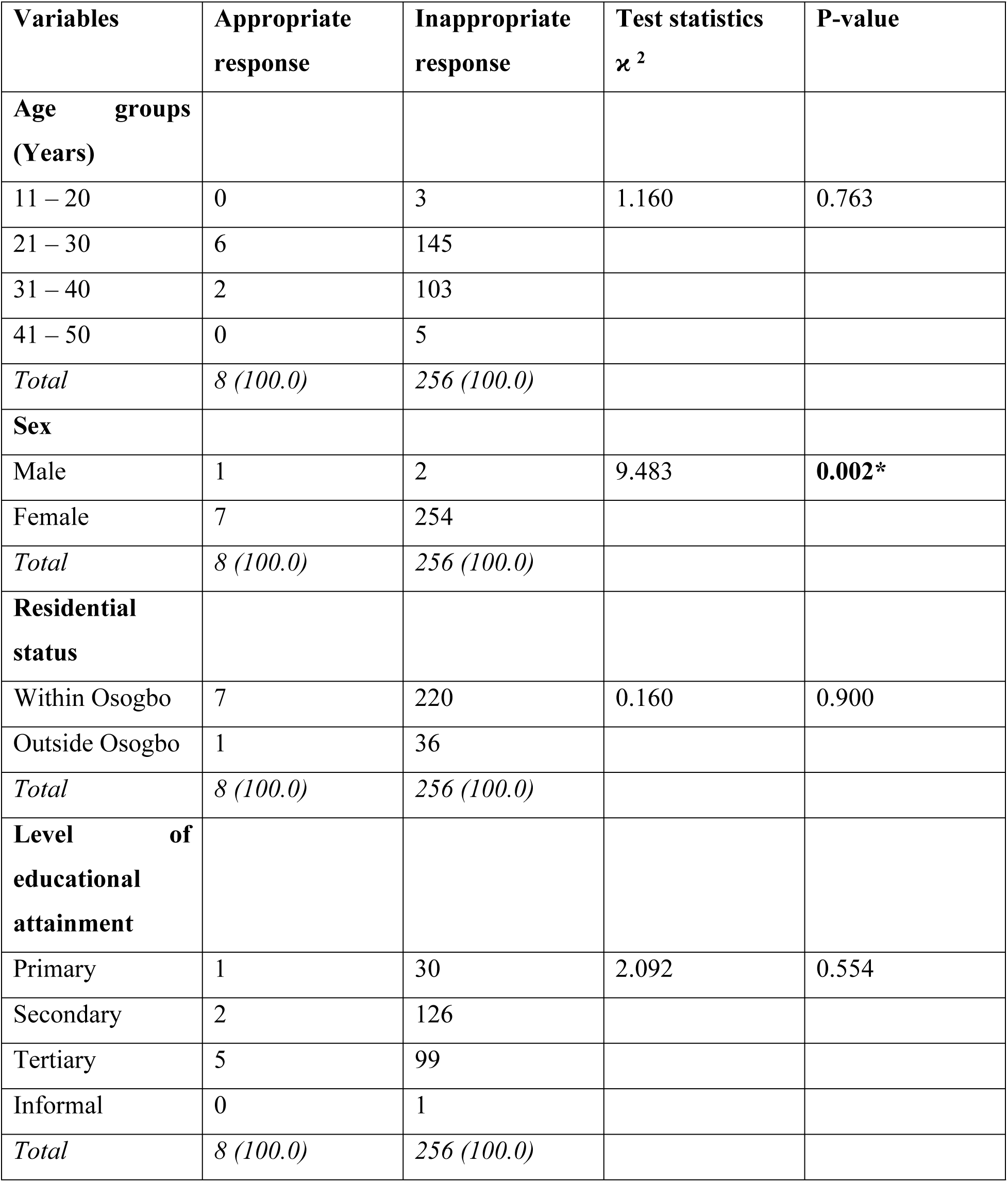
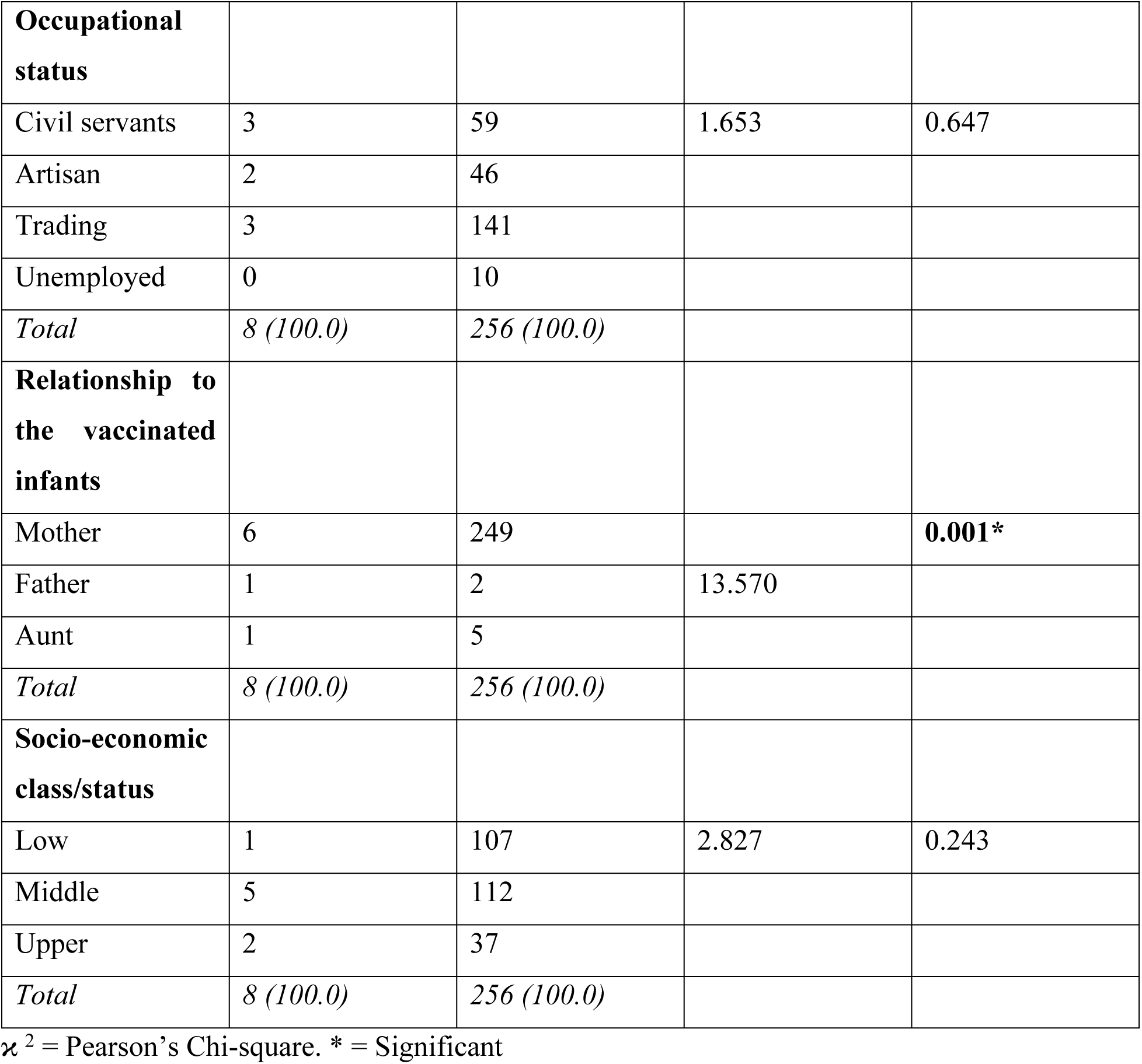
Determinants of Caregivers’ Response to the Occurrence of AEFI.

| **Variables** | **Appropriate response** | **Inappropriate response** | **Test statistics $\chi^2$** | **P-value** |
| --- | --- | --- | --- | --- |
| **Age groups (Years)** |  |  |  |  |
| 11 – 20 | 0 | 3 | 1.160 | 0.763 |
| 21 – 30 | 6 | 145 |  |  |
| 31 – 40 | 2 | 103 |  |  |
| 41 – 50 | 0 | 5 |  |  |
| *Total* | *8 (100.0)* | *256 (100.0)* |  |  |
| **Sex** |  |  |  |  |
| Male | 1 | 2 | 9.483 | **0.002\*** |
| Female | 7 | 254 |  |  |
| *Total* | *8 (100.0)* | *256 (100.0)* |  |  |
| **Residential status** |  |  |  |  |
| Within Osogbo | 7 | 220 | 0.160 | 0.900 |
| Outside Osogbo | 1 | 36 |  |  |
| *Total* | *8 (100.0)* | *256 (100.0)* |  |  |
| **Level of educational attainment** |  |  |  |  |
| Primary | 1 | 30 | 2.092 | 0.554 |
| Secondary | 2 | 126 |  |  |
| Tertiary | 5 | 99 |  |  |
| Informal | 0 | 1 |  |  |
| *Total* | *8 (100.0)* | *256 (100.0)* |  |  |
| **Occupational status** |  |  |  |  |
| Civil servants | 3 | 59 | 1.653 | 0.647 |
| Artisan | 2 | 46 |  |  |
| Trading | 3 | 141 |  |  |
| Unemployed | 0 | 10 |  |  |
| *Total* | *8 (100.0)* | *256 (100.0)* |  |  |
| **Relationship to the vaccinated infants** |  |  |  |  |
| Mother | 6 | 249 |  | **0.001\*** |
| Father | 1 | 2 | 13.570 |  |
| Aunt | 1 | 5 |  |  |
| *Total* | *8 (100.0)* | *256 (100.0)* |  |  |
| **Socio-economic class/status** |  |  |  |  |
| Low | 1 | 107 | 2.827 | 0.243 |
| Middle | 5 | 112 |  |  |
| Upper | 2 | 37 |  |  |
| *Total* | *8 (100.0)* | *256 (100.0)* |  |  |

$\chi^2$ = Pearson's Chi-square. \* = Significant

Further statistical analysis using binomial regression revealed that being a mother as a relationship to the vaccinated infant was observed to be a significant determinant of caregivers’ response to occurrence of AEFI (AOR: 47.667, 95% CI: 2.977 – 763.339, p = 0.006) while sex of the caregivers played no significant determinant (AOR: 1.368, 95% CI: 0.041 – 45.592, p = 0.861).

## DISCUSSION

### Summary of principal findings

This study was carried out to determine the prevalence, pattern, and determinants of caregivers’ response regarding AEFI in selected hospitals in Osogbo, Southwest Nigeria. The prevalence of AEFI was 71.7%, with inconsolable crying/irritability, fever, and swelling at the vaccinated sites as the leading cases. Age of the infant, their weight, and gestational status at birth were predictors of the occurrence of AEFI, and the implicated vaccine schedules were at birth, 6^th^, 10^th^, and 14^th^ weeks. Only 3% of the caregivers presented at a healthcare facility when AEFI was noticed (appropriate response), and the relationship of the caregiver to the vaccinated infant (mother) was found to be a determinant of caregivers’ response to the occurrence of AEFI.

### Comparison with previous literature

The prevalence of AEFI observed in this study was higher than 43.5% 16 and 66.5% 17 reported by Lawan et al in Kano, Northern Nigeria, and by Ekwueme in Enugu, Southeast Nigeria, respectively. This variation may be due to differences in the vaccines available to the infants. Haemophilus influenzae type B vaccine (Hib), Pneumococcal conjugate vaccine (PCV), and Rotavirus vaccine were additional vaccines administered to the infants in this study compared with those of Lawan et al and Ekwueme. It was also appreciably higher than 11.9%, 18 19.0%, 19 22.7%, 20 and 32% 21 reported by researchers across India, Iran, Spain, and Poland, respectively. These differences may reflect ethnic and genetic variation among the infants, as well as differences in vaccine manufacturers across the regions.

Regarding the pattern of AEFI, irritability/persistent excessive crying was the most common AEFI. This finding contrasted with the result obtained by Aderibigbe et al. in a study from Ilorin, Nigeria. 22 The pattern obtained was also at variance with the findings of some researchers who documented fever as the most common AEFI. ^23, 24^. The reason for these differences might be that irritability/persistent excessive crying appears to be more noticeable to caregivers than fever. While the present study focused on all vaccines, Orji’s work focused only on events following measles immunization, and Puliyel’s work focused on events following pentavalent vaccination. These differences might be the reason for the dissimilarity in the pattern obtained.

The largest percentage of occurrence of AEFI in this study was associated with the at-birth, 6th, 10th, and 14th week schedules, while the vaccine combinations of note included pentavalent-based and BCG-based vaccines. This finding is corroborated by findings from previous studies. ^18, 22, 25^. Interestingly, the occurrence of AEFI was highest with the first dose of Pentavalent-based vaccines, and it decreased with the subsequent doses. This observation has also been documented in some literature. ^18, 26^

Age of the infants was observed to be a predictor of occurrence of AEFI in this study; this finding is consistent with what Adam et al reported in Benin City, Nigeria.^25^ Outside the shores of Nigeria, Haritha et al reported a similar observation in India.^27^ The consistency in these results is likely to be due to relative similarity in the sociodemographic characteristics of these infants as well as the similarity in the methodology used in carrying out this study. Weight was also found to be an important predictor of AEFI in this study and was in tandem with the findings of Sebastian et al in an Indian study. ^26^ Concerning the gestational status of the infants at the time of birth, the present study noted that it was a predictor of the occurrence of AEFI, and this result was in line with the findings of Rodrigues Costa et al in a Brazilian study. ^28^

Despite the high occurrence of AEFI as recognized by the caregivers in this present study, it is surprising to note that only 3% of the caregivers exercised the appropriate response by reporting the events and presenting their infants for proper evaluation in the hospital. This finding differed from 0.0% to 38.4% reported in other literature ^13, 25, 29–31^. The reason for the variance in the present study might be differences in the definition of an appropriate response used in these studies. The relationship of the caregivers to the vaccinated infants was found to be a determinant of response to the occurrence of AEFI. Being a mother was a significant determinant, which was not a surprise given the physiological bond between mothers and their infants and mothers’ role in family health.^32^

### Implications for policy, practice, and future research

Arising from this study, it is obvious that the percentage of infants with AEFI is increasing compared to what other literature has documented. This has a far-reaching effect on immunization programmes, as vaccine hesitancy might be on the rise, and this calls for more public enlightenment to allay fear and correct misconceptions while reinforcing the importance of immunization. Most of the caregivers were able to identify what AEFI conditions are, and this portends a good omen for child health. The fact that inconsolable crying/irritability, fever, and swelling at the vaccination site were the leading AEFIs in this study suggests that caregivers, healthcare workers, and healthcare facilities should be well prepared and equipped with the necessary resources to manage these conditions. While some home care management may be encouraged, it is important to emphasize that an appropriate response to AEFI is to report to the healthcare facility for proper evaluation and management. This was not the case in the present study and, as such, calls for concern and the need for positive change to curtail the trend through greater public engagement, especially among caregivers. It is equally important to provide effective training and retraining for healthcare professionals on AEFI, particularly in identifying predictors of occurrence and determinants of caregivers’ response, as featured in the present study.

Knowledge about the vaccine schedules and the vaccines notorious for many occurrences of AEFI, as noted in the present study, would inform preparedness among the stakeholders (caregivers and healthcare professionals). It will also call for research into why the administration/receipt of these vaccines is often associated with many AEFI occurrences, with the possibility of finding definitive solutions, be it the development of new vaccines or ameliorating/attenuating the conditions of concern. Meaningful and productive collaboration is therefore encouraged by the government, non-governmental organizations, research funding agencies, the public, and healthcare professionals.

### Strengths and limitations of the study

Reliance on self-reported responses to questions about the occurrence of AEFI is likely to be plagued by social desirability bias and recall inaccuracies, which may lead to overestimation or underestimation of vital information. This was, however, addressed through a detailed explanation of the purpose of the study and the importance of giving correct responses to the questions asked. Also, absolute confidentiality of the caregivers’ responses was emphasized and upheld, and as such, potential concern of victimization or intimidation of any kind by virtue of their participation in the study was prevented. The cross-sectional design deployed in the conduct of this study is incapable of establishing causality, especially concerning the findings of AEFI predictors and the determinants of caregivers’ responses to the occurrence, thereby making it necessary to conclude using the findings obtained with caution.

The strength inherent in this study lies in the diverse population coverage from the multicentre participation of the caregivers, which to some extent allows for generalizability of the findings of the study.

### Conclusion

This study found a high prevalence of AEFI among infants in the studied Nigerian hospitals, with irritability/excessive crying, fever, and injection-site swelling being the most common. Infant age, weight, and gestational status at birth predicted AEFI occurrence, while the caregiver’s relationship to the infant—particularly being the mother—determined the response to AEFI. Despite this high burden, only 3.0% of caregivers who noticed AEFI presented to a healthcare facility; most self-medicated or took no action. These findings highlight the urgent need for targeted public enlightenment, caregiver education, and health-worker training on AEFI recognition, reporting, and appropriate management to improve vaccine safety and sustain confidence in immunization programmes.

## List of abbreviations

Abbreviation: Full term
AEFI: Adverse Events Following Immunization
AOR: Adjusted Odds Ratio
BCG: Bacillus Calmette-Guérin
CHEWS: Community Health Extension Workers
CI: Confidence Interval
CS: Caesarean Section
HBV: Hepatitis B Vaccine
Hib: Haemophilus influenzae type B vaccine
IPV: Inactivated Polio Vaccine *(not expanded in manuscript)*
OLFCH: Our Lady of Fatima Catholic Hospital
OPV: Oral Polio Vaccine
OSHREC: Osun State Health Research Ethical Committee
PCV: Pneumococcal Conjugate Vaccine
PHC(s): Primary Health Care Centre(s)
SD: Standard Deviation
SPSS: Statistical Package for Social Sciences
SSHA: State Specialists’ Hospital Asubiaro
SVD: Spontaneous Vertex Delivery
UNIOSUN: Osun State University
USA: United States of America
UTH: UNIOSUN Teaching Hospital
WHO: World Health Organization

## Declarations

### Ethics approval and consent to participate

This study was conducted in accordance with the ethical principles outlined in the Declaration of Helsinki. Ethical approval for this study was obtained from Osun State Health Research Ethical Committee with protocol number OSHREC/PRS/569T/1279. All ethical principles guiding the conduct of research such as informed consent, beneficence, non-maleficence, confidentiality, justice, autonomy etc., were strictly adhered to. Written informed consent was obtained from all the participants prior to participating in the study.

## Consent for publication

Not applicable.

## Acknowledgements

The authors sincerely appreciate all participants who took part in this study for their cooperation. Our big thanks also go the nurses at the immunization centres for their valuable guidance during data collection.

## Clinical trial number

Not applicable.

## Authors’ contributions

Olagunju, Adeyemo, and Odeyemi conceived and designed the study. Agbelegbe, Omoboyeje and Awodele acquired the data. Olagunju and Oyelami-Adegbite analysed and interpreted the data. Olagunju and Olabode drafted the manuscript. Agbelegbe, Adeyemo and Oninla critically revised the manuscript for important intellectual content. All authors approved the final version and agree to be accountable for all aspects of the work.

## Data availability

Data for this study is provided within the manuscript

